# Stratifying the risk of transition to adult-onset psychiatric disorders in adolescents with anxiety

**DOI:** 10.64898/2026.05.15.26353293

**Authors:** Charlotte A. Dennison, Amy Shakeshaft, Lucy Riglin, Frances Rice, Ole Andreassen, Helga Ask, Alexandra Havdahl, Daniel Pine, Joanna Martin, Anita Thapar

## Abstract

**Background:** Escalating mental health service demands have created a need to better identify young people most likely to require continued support from mental health services at the transition between childhood and adulthood. Anxiety is the most common adolescent mental health condition, yet its clinical significance and prognosis are not well understood. We aimed to examine the risk of young adult-onset psychiatric disorders in individuals with an adolescent anxiety disorder, and identify stratifiers of risk of subsequent psychiatric disorders in this group.

**Methods:** Individuals from the Norwegian Mother, Father, and Child Cohort Study (MoBa) with linked health records and aged 18 or over as of the 31^st^ December 2023 were included. Those diagnosed with any ICD-10 anxiety disorder when aged 10-17 years were defined as having an adolescent anxiety disorder (n=2107, controls n=47,582). Polygenic scores (PGS) for psychiatric and neurodevelopmental conditions were calculated using LDpred2. Anxiety, comorbidities, and parental psychiatric history were defined through linked ICD-10 diagnoses. Sex was defined through linked records. Individuals were defined as having a young adult-onset psychiatric disorder if they first received any new psychiatric diagnosis aged 18-24.

**Results:** Adolescent anxiety diagnosis was associated with increased risk of all adult-onset psychiatric disorders (HR= 2.33-8.65). Post-traumatic stress disorder PGS, parental history of severe mental illness, and female sex were associated with increased risk of transition to a young adult-onset psychiatric disorder in people with an adolescent anxiety disorder.

**Conclusions:** Adolescent anxiety greatly increases the risk of a psychiatric disorder during the transition to adult life. Clinicians should consider female sex and parental psychiatric history when prioritising young people with anxiety for adult mental health service support. Future research needs to further consider whether polygenic scores would aid risk stratification in clinical practice.

## Introduction

Demands on both child and adolescent (CAMHS) and adult mental health services (AMHS) have escalated globally in the last decade, especially after the Covid-19 pandemic^1,2^. Most AMHS users have previously been in contact with CAMHS^3^; yet moving across the two services is frequently difficult, resulting in service disengagement and treatment discontinuation^4^. Indeed, evidence suggests an increase in untreated cases of psychiatric disorder during the transition to adulthood, compared to during adolescence^5^.

Anxiety disorders, the most common paediatric mental health conditions^6^, are rarely associated with continued treatment in AMHS^4,7^. Many adolescents with anxiety show remission, and thus some clinicians view it as less severe or risky than psychiatric conditions such as psychosis, where referral to AMHS is more common^8,9^. However, adolescent anxiety is associated with various subsequent psychiatric problems, including depression, bipolar disorder, substance misuse, psychotic experiences, self-harm, and suicidal behaviours^10–15^. The combination of high prevalence and variable prognosis creates a major need to stratify adolescent anxiety disorders according to risk of subsequent psychiatric disorders. Evidence informing stratification would aid clinicians in prioritising treatment continuation for those at greatest need. However, there is limited research to stratify common psychiatric disorders, like anxiety, in adolescence.

Longitudinal studies suggest that severity and comorbidity among anxiety disorders increase risk of poorer psychiatric outcomes^16,17^. Although there is growing interest as to the potential clinical applications of genetic data in psychiatry^18^, so far, no studies have examined the extent to which psychiatric polygenic scores and parental psychiatric history predict new-onset psychiatric disorders in those with adolescent anxiety. Risk stratification has been highlighted as a promising area for clinical translation of genetic findings^19,20^. The increasing rates of both adolescent anxiety^21^ and AMHS referrals suggest that genetic information could be a valuable stratifier in this context.

In this study, we utilised a large Norwegian cohort linked to secondary healthcare data. We first characterised the risk of young adult (ages 18-24 years) psychiatric disorders in those with a recorded diagnosis of any adolescent anxiety disorder (ages 10-17 years). Second, we examined the contribution of psychiatric polygenic scores, parental psychiatric history, and clinical and demographic factors, to the risk of new young adult-onset psychiatric disorders in individuals with adolescent anxiety. Third, we examined whether effects are specific to particular anxiety disorders or outcomes, and examined sex differences, as prior research of adolescent depression suggests female sex is associated with worse adult mental health outcomes^22^.

## Methods

### Participants

Data were ascertained from the Norwegian Mother, Father and Child Cohort Study (MoBa)^23–25^. MoBa is a population-based pregnancy cohort study conducted by the Norwegian Institute of Public Health. Participants were recruited from all over Norway between 1999-2008. The women consented to participation in 41% of the pregnancies. The cohort includes approximately 114,500 children, 95,200 mothers and 75,200 fathers. The establishment of MoBa and initial data collection were based on a license from the Norwegian Data Protection Agency and approval from The Regional Committees for Medical and Health Research Ethics. The MoBa cohort is regulated by the Norwegian Health Registry Act. The current study was approved by The Regional Committees for Medical and Health Research Ethics (2016/1702). All procedures contributing to this work comply with the ethical standards of the relevant national and institutional committees on human experimentation and with the Helsinki Declaration of 1975, as revised in 2008. Blood samples were obtained from both parents during pregnancy and from mothers and children (umbilical cord) at birth^24^. For MoBa genotyping information, including quality control, see Corfield et al.^26^

Data were linked to the Norwegian Patient Registry (NPR), providing health records from secondary care services from 2008 up to 31^st^ December 2023^27^. The primary sample used for this study comprised all participants who had reached at minimum age 18 years by the time of the NPR cut-off (birth year ≤2005), filtered to exclude younger siblings. The maximum age was 24 years.

### Outcomes: Psychiatric disorders first diagnosed in young adulthood

ICD-10 diagnoses were used to determine age at diagnosis of the following psychiatric disorders that are common in young adults: i) psychotic disorder (F20-25, F28, F29), ii) bipolar disorder (F30-31), iii) major depressive disorder (MDD) (F32-33), iv) obsessive-compulsive disorder (OCD) (F42), v) post-traumatic stress disorder (PTSD) (F431), vi) eating disorder (F50), and vii) personality disorder (F60). Our primary outcome was any young adult-onset psychiatric disorder, defined as individuals who first received a diagnosis of any of these seven disorder types at age 18 years or over. Our secondary outcomes were the specific disorders listed above, diagnosed for the first time aged 18 or over. Where an individual received multiple diagnoses within a category, the earliest record determined age at first diagnosis. For example, a diagnosis of a depressive episode at age 16, followed by recurrent depressive disorder at age 19, would not be defined as an adult-onset disorder because the diagnoses are within the same category (MDD). We focused only on the onset of a new diagnosis category aged ≥18 (such as bipolar disorder in this example). We did not consider continuation of anxiety disorders into adulthood because our primary research question considered the onset of a different disorder in adulthood and because adult anxiety is typically managed under primary care, which is not captured by NPR.

### Exposure: Adolescent anxiety diagnosis

We defined our exposure group as anyone with a recorded diagnosis of any ICD-10 anxiety disorder (F40-F41, F93.0-93.2) during adolescence (aged 10-17). Adolescence was defined as starting from age 10, in accordance with the World Health Organisation definition. Our non-exposed group were individuals with no record of an anxiety disorder between ages 10-17. Individuals with an anxiety diagnosis recorded before age 10 and no record during adolescence were defined as ‘non-exposed’, whilst those first diagnosed prior to age 10 with further records of a diagnosis during adolescence were defined as ‘exposed’. Individual anxiety disorders were defined using the following ICD-10 codes based on the NPR: separation anxiety (F93.0), agoraphobia (F40.0), social anxiety (F40.1, F93.2), phobia (F40.2, F40.8, F40.9), panic disorder (F41.0), generalised anxiety disorder (F41.1), mixed anxiety disorder (F41.2, F41.3), and other anxiety disorder (F41.8, F41.9).

### Risk stratifiers: Psychiatric parental history and polygenic risk

Parental history of any psychiatric disorder was defined as the presence of any of the psychiatric disorders listed above in the mother or father. Parental history of severe mental illness (SMI) was defined as mother’s or father’s record of psychotic or bipolar disorder. Parental history of depression or anxiety was defined as a mother’s or father’s record of MDD or anxiety.

We used LDpred2^28^ and the largest and most recently published GWAS to calculate polygenic scores (PGS) for the following disorders: ADHD^29^, anxiety^30^, bipolar disorder^31^, MDD^32^, OCD^32^, PTSD^33^, and schizophrenia^34^. For further information, see Supplementary Method.

### Risk stratifiers: Demographic and clinical factors

Biological sex at birth was defined by birth records from the Medical Birth Registry of Norway, or by parent-report at 6 months of age when birth records were unavailable.

Psychiatric comorbidity was defined as having any of the aforementioned psychiatric diagnoses recorded between ages 10-17.

Neurodevelopmental comorbidity was defined as having any of the following diagnoses with onset any age prior to 18 years: i) ADHD (F90), ii) autism spectrum disorder (ASD, F84), iii) speech and language disorder (F80), iv) tic disorder (F95), v) developmental coordination disorder (F82), and vi) specific learning disorders (F81).

For data availability, see Supplementary Method.

## Statistical analysis

All analyses were conducted using R.

First, we used Cox regression, using the ‘survival’ package^35^, to examine the risk of any new adult-onset psychiatric disorder (≥18 years) in those with and without an adolescent anxiety disorder (10-17 years).

Second, we used Cox regression to examine stratifiers of risk for any adult-onset psychiatric disorder. Using univariate models, we tested for associations between any adult-onset psychiatric disorder and i) parental psychiatric history, ii) neurodevelopmental and psychiatric comorbidity, iii) sex, and iv) psychiatric and neurodevelopmental PGS. We examined the association between potential risk stratifiers and young adult-onset psychiatric disorders at a population level, regardless of adolescent anxiety diagnosis. Next, we included only individuals with a diagnosed anxiety disorder in adolescence. Then, we examined the interaction between risk stratifiers and adolescent anxiety to test whether effects differed by adolescent anxiety status.

Finally, we examined the absolute risk of developing an adult-onset psychiatric disorder in those with an adolescent-onset anxiety disorder, stratified by PGS and parental psychiatric history. Absolute risk was examined using the package ‘riskRegression’^36^. We stratified those with a youth anxiety disorder according to PGS percentile and parental history of SMI and examined the absolute risk of a young adult-onset psychiatric disorder. Individuals were first grouped according to whether they had an adolescent anxiety disorder and low (<90th centile) or high (>90th centile) PGS, and secondly by whether they had an adolescent anxiety disorder and parental history of SMI.

Primary results are corrected for multiple testing using false discovery rate (FDR), at a threshold of 0.05.

### Secondary analysis

To test whether observed effects were specific to anxiety disorder subtypes, we performed Cox regression analyses between each anxiety disorder category and any adult-onset psychiatric disorder.

To examine whether adolescent anxiety was associated with specific adult-onset psychiatric disorders, we repeated the primary Cox regression analyses with each adult psychiatric disorder as the outcome. To test for sex-specific effects, we repeated the primary analyses stratified by sex.

## Results

### Descriptives

There were 49,689 unrelated participants aged 18-24 by the end of the study period. Of these, 2,107 people (4.2%) had a diagnosed anxiety disorder during adolescence. Of those with an adolescent anxiety disorder, 1478 (70.1%) were female, and the mean age at anxiety diagnosis was 14.8 years. The number of individuals with each type of anxiety disorder is presented in Table S1. Social anxiety was the most common anxiety disorder, occurring in 1.7% of adolescents.

2,109 (4.4%) of all individuals developed a young adult-onset psychiatric disorder by the end of the study period, of whom 1,501 (71.2%) were female. The number of individuals with each young adult psychiatric disorder is presented in Table 1. Sample sizes for the potential risk stratifiers are presented in Table S2.

**Table 1.**
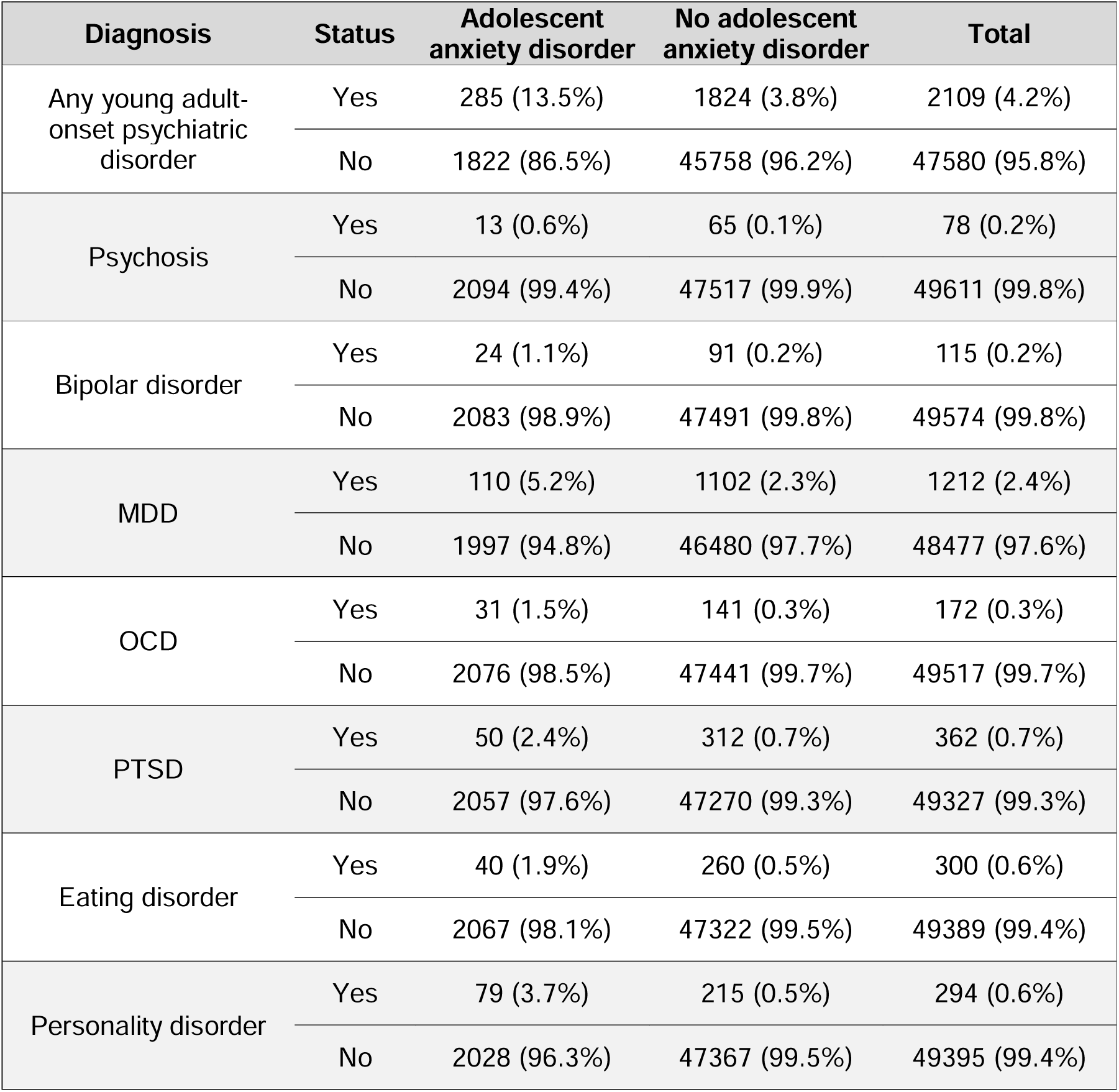
Frequency of young adult-onset psychiatric disorder, stratified by those with and without an adolescent anxiety disorder. Percentages represent column proportions.

| Diagnosis | Status | Adolescent anxiety disorder | No adolescent anxiety disorder | Total |
| --- | --- | --- | --- | --- |
| Any young adult-onset psychiatric disorder | Yes | 285 (13.5%) | 1824 (3.8%) | 2109 (4.2%) |
|  | No | 1822 (86.5%) | 45758 (96.2%) | 47580 (95.8%) |
| Psychosis | Yes | 13 (0.6%) | 65 (0.1%) | 78 (0.2%) |
|  | No | 2094 (99.4%) | 47517 (99.9%) | 49611 (99.8%) |
| Bipolar disorder | Yes | 24 (1.1%) | 91 (0.2%) | 115 (0.2%) |
|  | No | 2083 (98.9%) | 47491 (99.8%) | 49574 (99.8%) |
| MDD | Yes | 110 (5.2%) | 1102 (2.3%) | 1212 (2.4%) |
|  | No | 1997 (94.8%) | 46480 (97.7%) | 48477 (97.6%) |
| OCD | Yes | 31 (1.5%) | 141 (0.3%) | 172 (0.3%) |
|  | No | 2076 (98.5%) | 47441 (99.7%) | 49517 (99.7%) |
| PTSD | Yes | 50 (2.4%) | 312 (0.7%) | 362 (0.7%) |
|  | No | 2057 (97.6%) | 47270 (99.3%) | 49327 (99.3%) |
| Eating disorder | Yes | 40 (1.9%) | 260 (0.5%) | 300 (0.6%) |
|  | No | 2067 (98.1%) | 47322 (99.5%) | 49389 (99.4%) |
| Personality disorder | Yes | 79 (3.7%) | 215 (0.5%) | 294 (0.6%) |
|  | No | 2028 (96.3%) | 47367 (99.5%) | 49395 (99.4%) |

**Table 2.** Results from univariate Cox regression analyses of the development of a young adult-onset psychiatric disorder in those with adolescent anxiety disorder. P-values are uncorrected for multiple comparisons, whilst Q-values are corrected at a threshold of p<0.05.

| Risk stratifiers | Hazard ratio | 95% CI |  | P-value | Q-value |
| --- | --- | --- | --- | --- | --- |
|  |  | Lower | Upper |  |  |
| ***Polygenic scores*** |  |  |  |  |  |
| ADHD PGS | 1.17 | 1.02 | 1.35 | 0.028 | 0.11 |
| Anxiety PGS | 1.09 | 0.94 | 1.26 | 0.24 | 0.32 |
| Bipolar disorder PGS | 1.11 | 0.96 | 1.28 | 0.15 | 0.24 |
| MDD PGS | 1.11 | 0.97 | 1.28 | 0.14 | 0.24 |
| OCD PGS | 1.13 | 0.98 | 1.31 | 0.084 | 0.22 |
| PTSD PGS | 1.24 | 1.07 | 1.43 | 3.9E-03 | 0.031 |
| Schizophrenia PGS | 1.03 | 0.90 | 1.18 | 0.65 | 0.65 |
| ***Parental history*** |  |  |  |  |  |
| Parental history of any psychiatric disorder | 1.05 | 0.83 | 1.34 | 0.66 | 0.80 |
| Parental history of SMI | 1.94 | 1.27 | 2.94 | 2.0E-03 | 0.006 |
| Parental history of anxiety or depression | 0.97 | 0.75 | 1.25 | 0.81 | 0.81 |
| ***Demographic and clinical factors*** |  |  |  |  |  |
| Psychiatric co-morbidity | 1.29 | 1.02 | 1.63 | 0.035 | 0.071 |
| Co-occurring neurodevelopmental condition | 0.89 | 0.60 | 1.32 | 0.56 | 0.80 |
| Female sex | 2.38 | 1.73 | 3.26 | 7.7E-08 | 4.6E-07 |

### Association between adolescent anxiety and young adult-onset psychiatric disorders

Individuals with adolescent anxiety had a 3.80 times elevated risk of developing any young adult-onset psychiatric disorder compared to those without adolescent anxiety (95%CI=3.35-4.30, q=1.4x10^-96^). All types of anxiety disorder were associated with increased likelihood of any adult-onset disorder, with the strongest effect observed for panic disorder (Table S3, HR range=2.74-4.63).

When running separate models for each adult psychiatric disorder category, the presence of any adolescent anxiety disorder was associated with every young adult psychiatric disorder; most strongly with personality disorder (HR=8.65 [6.69-11.20], q=7.6x10^-60^); see Figure 1 and Table S4.

**Figure 1.**
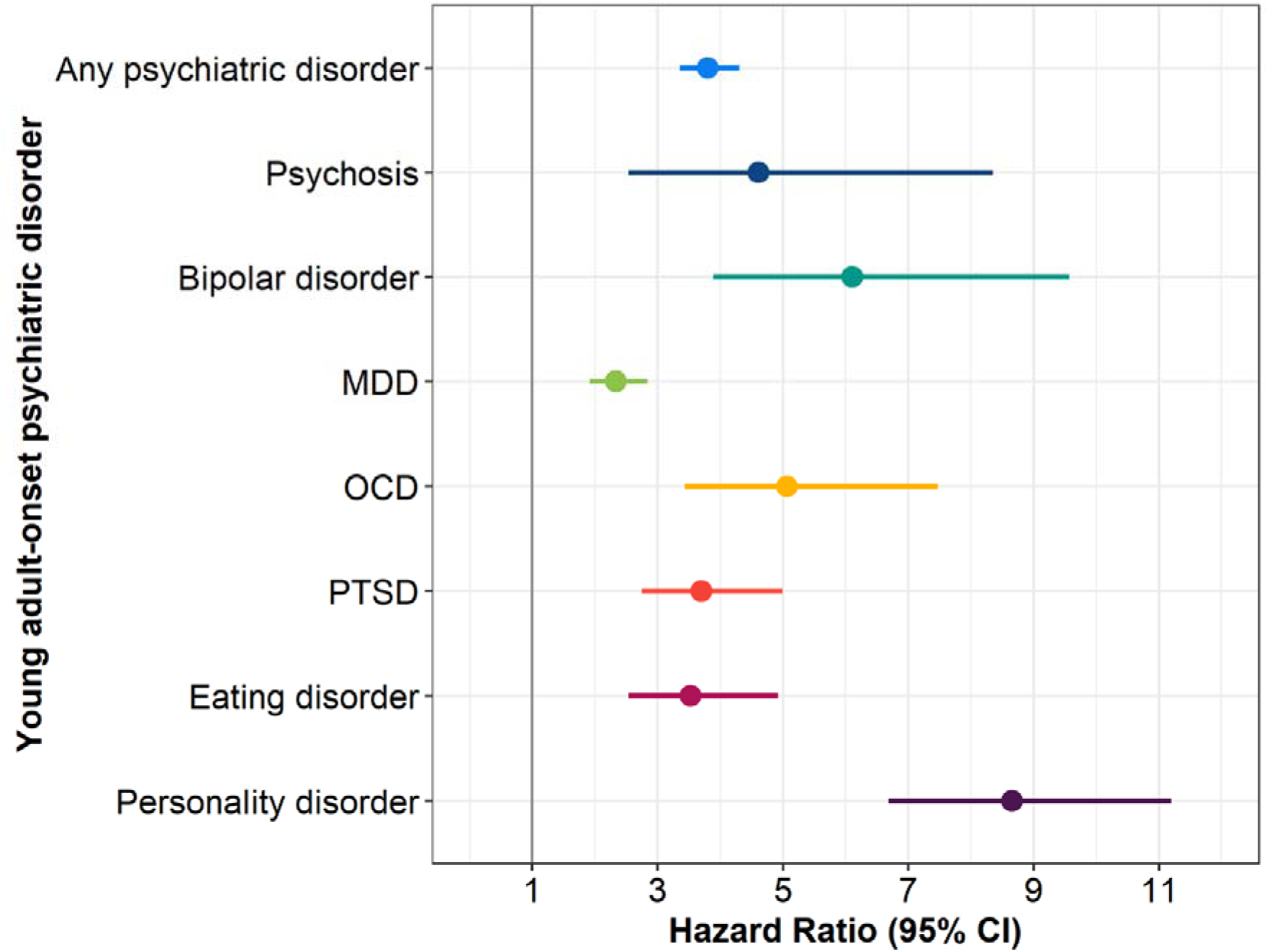
Hazard ratios and 95% confidence intervals of the association between adolescent anxiety and young adult-onset psychiatric disorders. MDD – major depressive disorder. OCD – obsessive compulsive disorder. PTSD – post-traumatic stress disorder

The likelihood of developing any psychiatric disorder following adolescent anxiety did not differ between males and females (Interaction: HR=0.97[0.79-1.35, p=0.87) (Table S5).

### Risk stratifiers for transitioning from adolescent anxiety disorder to young adult-onset psychiatric disorder

In the full sample, all stratifiers, except OCD PGS, were associated with increased risk of any young adult-onset psychiatric disorder (Table S6).

In individuals with adolescent anxiety, only PTSD PGS, parental history of SMI, and female sex were associated with risk of young adult-onset psychiatric disorder (Table 1, Figure 2). ADHD PGS and psychiatric comorbidity showed weaker evidence of association that did not survive correction for multiple testing. Interaction analysis did not support a difference in the strength of association between risk stratifiers and any young adult-onset disorder by adolescent anxiety status (Table S6). Associations between PGS and each separate psychiatric disorder are shown in Table S7.

**Figure 2.**
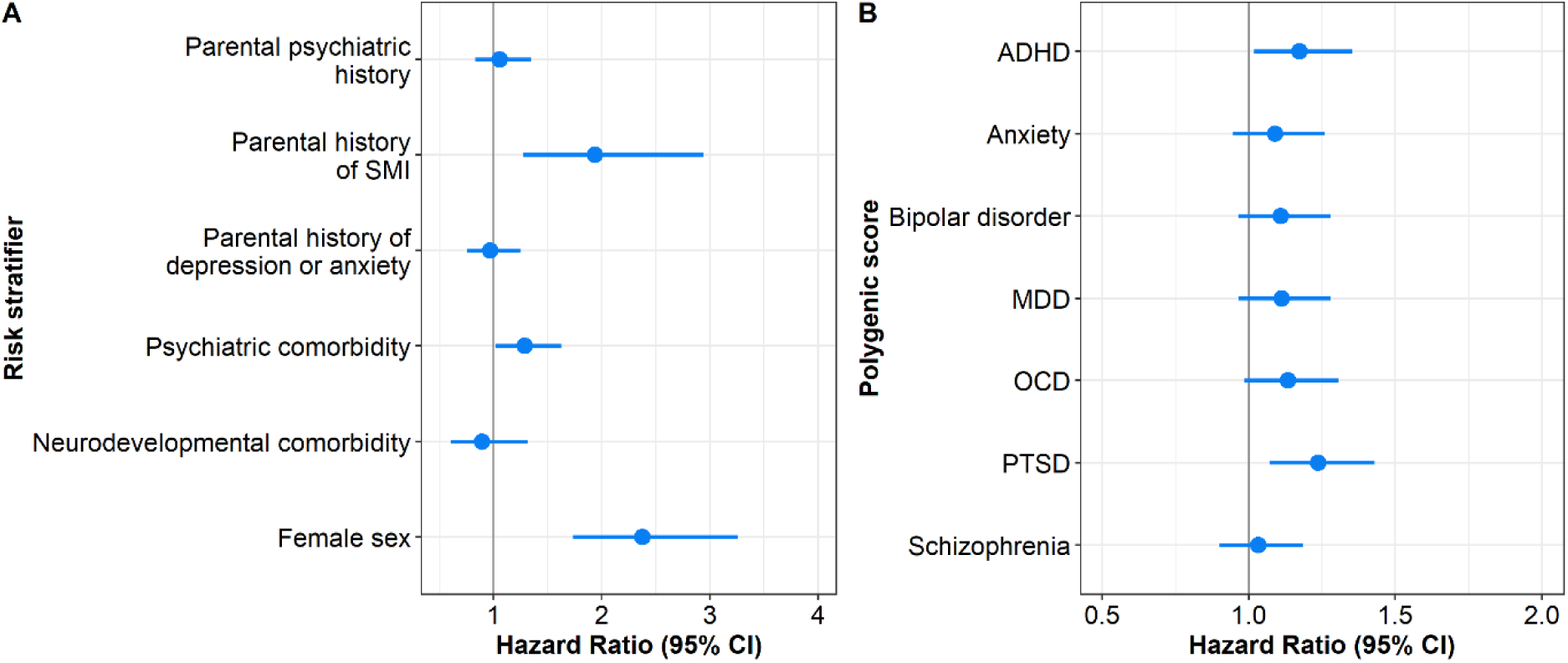
Hazard ratios and 95% confidence intervals for associations between risk stratifiers and any young adult-onset psychiatric disorder in individuals with adolescent anxiety disorder. A) demographic, clinical and parental history risk stratifiers, B) polygenic scores. ADHD – attention deficit hyperactivity disorder. MDD – major depressive disorder. OCD – obsessive compulsive disorder. PTSD – post-traumatic stress disorder.

In males with adolescent anxiety, ADHD PGS, OCD PGS, schizophrenia PGS, and psychiatric comorbidity were also associated with adult-onset disorder. Interaction analyses supported evidence of sex differences in the association between any young adult-onset psychiatric disorder and OCD PGS and psychiatric comorbidity (Table S5).

### Absolute risk of young adult-onset psychiatric disorder: the contribution of psychiatric PGS and parental history

The risk of developing any young adult-onset psychiatric disorder before age 25 was 4.8% (95% CI=4.53-5.08%) in people without adolescent anxiety, and 17.05% (95% CI=15.2-18.91%) in people with adolescent anxiety (Figure S1). Figure 3 displays the absolute risk of a psychiatric disorder by adolescent anxiety status and A) PTSD PGS and B) parental history of SMI. The absolute risk over time for each group by the other PGS is displayed in Figure S2. When stratifying by PTSD PGS, the highest absolute risk of any young adult-onset psychiatric disorder was seen in those with adolescent anxiety and high PTSD PGS, with an absolute risk of 24.0% (95% CI= 17.7-30.4%) before age 25. Individuals without anxiety with low PTSD PGS had an absolute risk of any young adult-onset psychiatric disorder of 4.5% (CI=4.1-4.8%). When stratifying by parental SMI history, absolute risk of any young adult-diagnosed psychiatric disorder was highest in individuals with adolescent anxiety and parental history of SMI, at 29.4% (CI=19.5-39.2%), compared to 4.7% (CI=4.4-4.9%) in individuals without adolescent anxiety and no parental history of SMI. We could not stratify by PGS and parental SMI combined due to low sample size.

**Figure 3.**
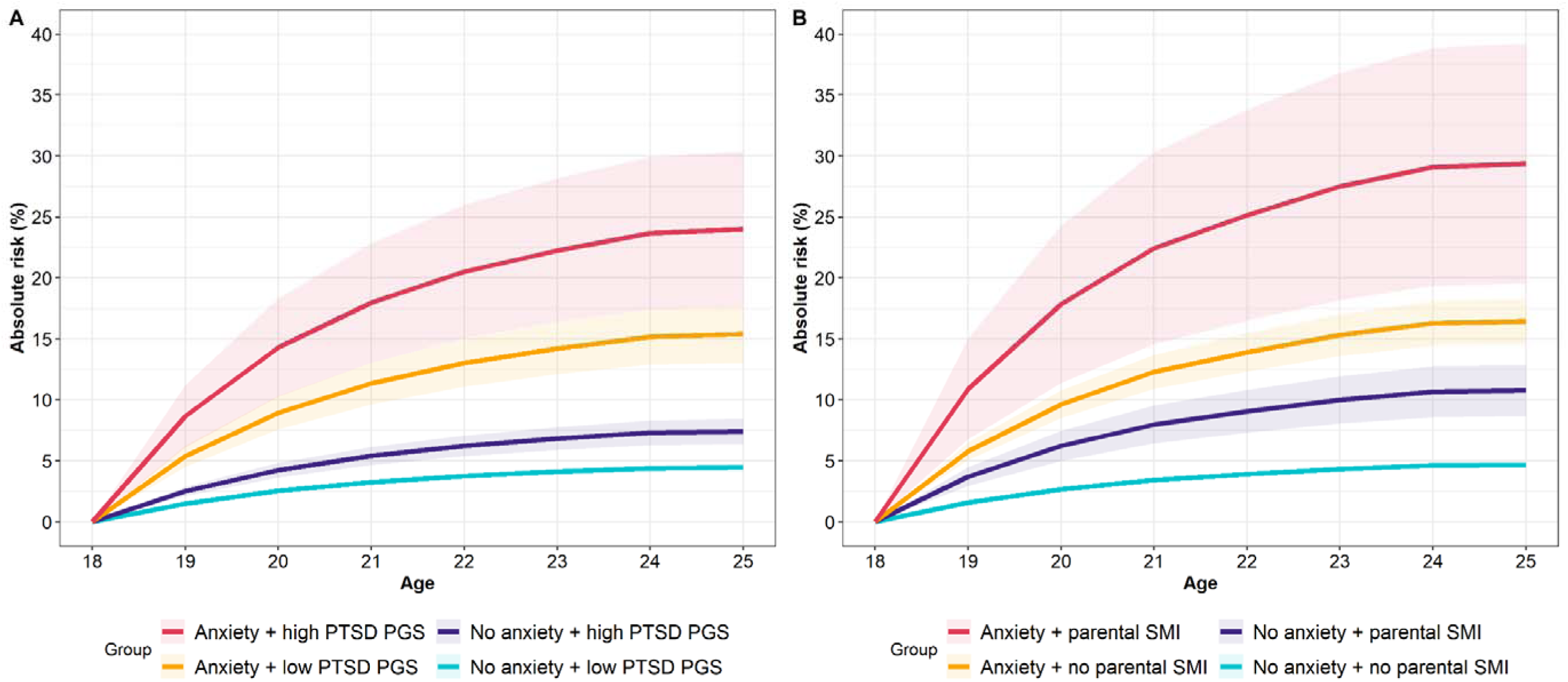
Absolute risk of developing any adult-onset psychiatric disorder in individuals with and without adolescent anxiety, stratified by A) High (>90^th^ percentile) and low (<90^th^ percentile) PTSD PGS, and B) parental history of SMI. Solid lines refer to the estimate of absolute risk and shaded areas represent 95% confidence intervals.

## Discussion

We examined the risk of psychiatric disorders first diagnosed in young adulthood among those with an adolescent-onset anxiety disorder. Adolescent anxiety was associated with an increased risk of all young adult-onset psychiatric disorders (≥18 years), with the most elevated risk observed for personality disorders^10–13,15^. An elevated risk was observed for every type of anxiety disorder. In those with adolescent anxiety, PTSD PGS, parental history of SMI, and female sex were associated with increased risk of transition to a young adult-onset psychiatric disorder. We did not identify evidence to suggest that baseline psychiatric or neurodevelopmental comorbidity was associated with additional risk.

Our findings highlight that individuals with adolescent anxiety disorders are at higher risk of transition to adult psychiatric disorder, requiring specialist support. We found that PTSD PGS, parental history of SMI and being female were risk markers, which could help stratify those at highest risk of developing a psychiatric disorder in young adulthood. It is intriguing that PTSD polygenic score also predicted transition to adult disorder; given their novelty, these findings require replication. Adverse childhood experiences are a known risk factor for adolescent anxiety^37^, and our data cannot address the possible misdiagnosis of anxiety among young people with underlying PTSD. The PTSD GWAS may also capture genetic association with trauma exposure, i.e. gene-environment correlation, which becomes more important at the transition to adult life and may be important for explaining patterns of persistence in mental health problems over time^38^. Surprisingly, we did not observe strong evidence of an association between adolescent comorbidity and young adult-onset psychiatric disorders. By comparison, in the full sample, both psychiatric and neurodevelopmental comorbidity were markers of risk for adult psychiatric disorders. 37.4% of individuals with anxiety had a mental health comorbidity in adolescence. Therefore it may be that comorbidity is too common or heterogenous in adolescents with anxiety to be a useful stratifier of outcome.

There is growing interest in the potential uses of genomic data in clinical psychiatry for stratifying disorders^18,20^. Using PGS to evaluate prognosis and guide clinical care via precision medicine has been highlighted as the most likely future implementation^18^. In our study, among individuals with adolescent anxiety, having a parental history of SMI yielded an absolute risk of 29.4% for psychiatric disorder by age 25, which was almost double the risk observed in individuals with adolescent anxiety and no parental history of SMI (16.4%). Whilst there is interest in developing formal risk prediction models, these are complex to implement in practice^39^, and risk indicators that are simple and easy to assess are extremely valuable. Combining PGS with parental psychiatric history has also been shown to predict risk of MDD^19^. However, a limitation of our study is that we were unable to test the combined risk of high PGS and parental history of SMI on young adult-onset psychiatric disorders in individuals with adolescent anxiety due to low sample sizes.

Much of the existing literature on adolescent anxiety has relied on questionnaires^14^. Whilst these provide valuable information, they are limited in their clinical translation as most individuals with elevated anxiety symptoms will not reach secondary care services^40^.

A strength of our study was the use of healthcare records, linked to genomic data, which allowed us to focus specifically on those in contact with specialist services. As healthcare records are systematically collected, this alleviates issues of attrition typically found in longitudinal data. However, the study design also means we focused on those in secondary care, who have more severe disorders. Additionally, the first registration to NPR started in 2008, and parents with psychiatric records exclusively prior to this date will have been missed. We recognise that many common mental health problems for young people and adults are managed in primary care and that many affected individuals do not access specialist healthcare.

One limitation to our study is the limited age at follow-up of our sample. MoBa participants were recruited over a 10-year period and most individuals in our sample had only recently turned 18. Therefore, it is possible that individuals with adolescent anxiety may develop subsequent psychiatric disorders in the future, that we were unable to capture. Another weakness of our study is that almost all individuals are of European genetic ancestry, and it is unclear how our findings relate to individuals from other genetic ancestries. Furthermore, Norway is a high-income country with public healthcare, thus our findings may not be applicable to lower income countries or countries where healthcare is not freely available.

Overall, our findings demonstrate that adolescents with an anxiety disorder are at increased risk of developing a new, additional psychiatric disorder in early adulthood. Female sex and parental history of SMI predicted transition to adult psychiatric disorders within this group, as did PTSD PGS. Further research is needed to replicate these findings before evaluating how well risk stratification models would work in adolescent anxiety clinical services when planning transition to adult services.

## Acknowledgements

This work was supported by the Wolfson Centre for Young People’s Mental Health, established with support from the Wolfson Foundation. The Norwegian Mother, Father and Child Cohort Study is supported by the Norwegian Ministry of Health and Care Services and the Ministry of Education and Research. We are grateful to all the participating families in Norway who take part in this on-going cohort study. For generating high-quality genomic data, we thank the Norwegian Institute of Public Health (NIPH), the HARVEST collaboration, the NORMENT Centre at the University of Oslo, the Centre for Diabetes Research at the University of Bergen, deCODE Genetics, the Research Council of Norway, the SouthEastern and Western Norway Regional Health Authorities, the ERC AdG, Stiftelsen KG Jebsen, the Trond Mohn Foundation, and the Novo Nordisk Foundation.

Part of this work was performed on the TSD (Tjeneste for Sensitive Data) facilities, owned by the University of Oslo, operated and developed by the TSD service group at the University of Oslo, IT-Department (USIT). The analyses were also performed on resources provided by Sigma2 - the National Infrastructure for High-Performance Computing and Data Storage in Norway. Data from the Norwegian Patient Registry has been used in this publication. The interpretation and reporting of these data are the sole responsibility of the authors, and no endorsement by the Norwegian Patient Registry is intended nor should be inferred. JM was funded by the Welsh Government through Health and Care Research Wales via an NIHR Advanced Fellowship (Ref: NIHR-FS(A)-2022).

Daniel Pine, MD was supported by project ZIA-MH002782 from the Intramural Research programs (IRP) at the National Institute of Mental Health. As such, the research was supported [in part] by the IRP of the National Institutes of Health (NIH). The contributions of the NIH author(s) are considered Works of the United States Government. The findings and conclusions presented in this paper are those of the author(s) and do not necessarily reflect the views of the NIH or the U.S. Department of Health and Human Services.

HA was funded by NordForsk (156298 and 230738) and RCN (324620). OAA was supported by the RCN (324499, 324252, 296030) Nordforsk (164218) and South East Norway Health Authority (2023-031)

## Conflicts of interest

OAA is a consultant to Precision Health and has received speaker’s honorariums from BMS, Lilly, Lundbeck, Janssen, Otsuka, Sunovion. The remaining authors report no conflicts of interest.

## Data availability

Data from MoBa and the Medical Birth Registry of Norway used in this study are managed by the Norwegian Institute of Public Health, and can be made available to researchers upon approval from the Regional Committees for Medical and Health Research Ethics (REC), compliance with the EU General Data Protection Regulation (GDPR) and approval from the data owners. The consent given by the participants does not allow for storage of data on an individual level in repositories or journals. Researchers who want access to data sets for replication should apply through helsedata.no. Access to data sets requires approval from The Regional Committee for Medical and Health Research Ethics in Norway and an agreement with MoBa.

## Notes

### Competing Interest Statement

OAA is a consultant to Precision Health and has received speakers honorariums from BMS, Lilly, Lundbeck, Janssen, Otsuka, Sunovion. The remaining authors report no conflicts of interest.

### Funding Statement

This work was supported by the Wolfson Centre for Young Peoples Mental Health, established with support from the Wolfson Foundation. The Norwegian Mother, Father and Child Cohort Study is supported by the Norwegian Ministry of Health and Care Services and the Ministry of Education and Research. We are grateful to all the participating families in Norway who take part in this on-going cohort study. For generating high-quality genomic data, we thank the Norwegian Institute of Public Health (NIPH), the HARVEST collaboration, the NORMENT Centre at the University of Oslo, the Centre for Diabetes Research at the University of Bergen, deCODE Genetics, the Research Council of Norway, the SouthEastern and Western Norway Regional Health Authorities, the ERC AdG, Stiftelsen KG Jebsen, the Trond Mohn Foundation, and the Novo Nordisk Foundation.
Part of this work was performed on the TSD (Tjeneste for Sensitive Data) facilities, owned by the University of Oslo, operated and developed by the TSD service group at the University of Oslo, IT-Department (USIT). The analyses were also performed on resources provided by Sigma2 - the National Infrastructure for High-Performance Computing and Data Storage in Norway. Data from the Norwegian Patient Registry has been used in this publication. The interpretation and reporting of these data are the sole responsibility of the authors, and no endorsement by the Norwegian Patient Registry is intended nor should be inferred.
JM was funded by the Welsh Government through Health and Care Research Wales via an NIHR Advanced Fellowship (Ref: NIHR-FS(A)-2022).
Daniel Pine, MD was supported by project ZIA-MH002782 from the Intramural Research programs (IRP) at the National Institute of Mental Health. As such, the research was supported [in part] by the IRP of the National Institutes of Health (NIH). The contributions of the NIH author(s) are considered Works of the United States Government. The findings and conclusions presented in this paper are those of the author(s) and do not necessarily reflect the views of the NIH or the U.S. Department of Health and Human Services.
HA was funded by NordForsk (156298 and 230738) and RCN (324620). OAA was supported by the RCN (324499, 324252, 296030) Nordforsk (164218) and South East Norway Health Authority (2023-031)

### Author Declarations

The establishment of MoBa and initial data collection were based on a license from the Norwegian Data Protection Agency and approval from The Regional Committees for Medical and Health Research Ethics. The current study was approved by The Regional Committees for Medical and Health Research Ethics (2016/1702).

