## Supplementary Method for "Stratifying the risk of transition to adult-onset psychiatric disorders in adolescents with anxiety"

[**Table S4** Hazard ratios of the association between adolescent anxiety and diagnosis of specific psychiatric disorders in young adulthood (≥ 18 years). P-values are fdr-corrected at p<0.05.	6](#_Toc221698346)

### Supplementary Method

*Polygenic scores*

Polygenic scores (PGS) were calculated using the ‘genotools’ package in R (<https://github.com/psychgen/genotools>), which implements a quality control pipeline and calculates polygenic scores using LDpred2 (1) software. PGS were calculated using the largest available GWAS for ADHD(2), anxiety(3), bipolar disorder(4), MDD(5), OCD(6), PTSD(7), and schizophrenia(8). For GWAS including MoBa in the original sample, we used summary statistics that excluded MoBa. PGS were regressed on genotyping batch, imputation batch, and the first 20 principal components, derived previously by Corfield et al. (9). PGS were then standardised to enable cross-PGS comparisons.

*Data availability*

Data from the Norwegian Mother, Father and Child Cohort Study is managed by the Norwegian Institute of Public Health. Access requires approval from the Regional Committees for Medical and Health Research Ethics (REC), compliance with GDPR, and data owner approval. Participant consent does not allow individual-level data storage in repositories or journals. Researchers seeking access for replication must apply via www.helsedata.no.

### Supplementary Tables

#### **Table S1** Number of individuals with different types of adolescent anxiety. Percentages are calculated across rows.

NB: There were too few individuals with separation anxiety (F93.0) and an adult-onset psychiatric disorder to permit analysis of this group.

| **Anxiety type** | **Anxiety status** | **N without adult-onset disorder** | **N with adult-onset disorder** | **Total** |
| --- | --- | --- | --- | --- |
| Generalised anxiety disorder | Without | 47270 (95.8%) | 2067 (4.2%) | 49337 |
|  | With | 310 (88.1%) | 42 (11.9%) | 352 |
| Specific phobia | Without | 47298 (95.8%) | 2074 (4.2%) | 49372 |
|  | With | 282 (89%) | 35 (11%) | 317 |
| Agoraphobia | Without | 47481 (95.8%) | 2094 (4.2%) | 49575 |
|  | With | 99 (86.8%) | 15 (13.2%) | 114 |
| Panic disorder | Without | 47428 (95.8%) | 2077 (4.2%) | 49505 |
|  | With | 152 (82.6%) | 32 (17.4%) | 184 |
| Social anxiety | Without | 46886 (95.9%) | 1983 (4.1%) | 48869 |
|  | With | 694 (84.6%) | 126 (15.4%) | 820 |
| Mixed anxiety | Without | 47284 (95.8%) | 2056 (4.2%) | 49340 |
|  | With | 296 (84.8%) | 53 (15.2%) | 349 |
| Other anxiety | Without | 47304 (95.8%) | 2073 (4.2%) | 49377 |
|  | With | 276 (88.5%) | 36 (11.5%) | 312 |

#### **Table S2**. Number of individuals with adolescent anxiety with and without each stratifier. Percentages are calculated across rows.

SMI=severe mental illness.

| **Stratifier** | **Status** | **With anxiety + no adult-onset disorder** | **With anxiety + adult-onset disorder** |
| --- | --- | --- | --- |
| Sex | Male | 583 (92.7%) | 46 (7.3%) |
|  | Female | 1239 (83.8%) | 239 (16.2%) |
| Mental health comorbidity | Without | 1156 (87.7%) | 162 (12.3%) |
|  | With | 666 (84.4%) | 123 (15.6%) |
| Neurodevelopmental comorbidity | Without | 1621 (86.3%) | 257 (13.7%) |
|  | With | 201 (87.8%) | 28 (12.2%) |
| Parental psychiatric history (any) | Without | 1180 (86.7%) | 181 (13.3%) |
|  | With | 642 (86.1%) | 104 (13.9%) |
| Parental history (depression/anxiety) | Without | 1255 (86.3%) | 199 (13.7%) |
|  | With | 567 (86.8%) | 86 (13.2%) |
| Parental history (SMI) | Without | 1746 (87%) | 261 (13%) |
|  | With | 76 (76%) | 24 (24%) |

#### **Table S3** Hazard ratios of the association between specific types of adolescent anxiety and developing any adult-onset psychiatric disorder (≥18 years).

| **Anxiety type** | **Hazard Ratio** | **Lower CI** | **Upper CI** | **P-value** |
| --- | --- | --- | --- | --- |
| Generalised anxiety | 3.05 | 2.25 | 4.14 | 8.7E-13 |
| Specific phobia | 2.74 | 1.96 | 3.82 | 3.4E-09 |
| Agoraphobia | 3.32 | 2.00 | 5.52 | 3.6E-06 |
| Panic disorder | 4.63 | 3.27 | 6.57 | 7.6E-18 |
| Social anxiety | 4.16 | 3.47 | 4.98 | 3.1E-54 |
| Mixed anxiety | 3.93 | 2.99 | 5.16 | 8.0E-23 |
| Other anxiety | 2.89 | 2.08 | 4.01 | 2.9E-10 |

#### **Table S4** Hazard ratios of the association between adolescent anxiety and diagnosis of specific psychiatric disorders in young adulthood (≥ 18 years). Q-values are corrected at p<0.05.

| **Outcome diagnosis** | **Hazard Ratio** | **Lower CI** | **Upper CI** | **P-value** | **Q-value** |
| --- | --- | --- | --- | --- | --- |
| Psychosis | 4.61 | 2.54 | 8.35 | 5.0E-07 | 5.0E-07 |
| Bipolar disorder | 6.10 | 3.89 | 9.57 | 3.2E-15 | 4.3E-15 |
| MDD | 2.33 | 1.92 | 2.83 | 2.7E-17 | 5.5E-17 |
| OCD | 5.07 | 3.44 | 7.48 | 2.8E-16 | 4.5E-16 |
| PTSD | 3.70 | 2.75 | 4.99 | 8.7E-18 | 2.3E-17 |
| Eating disorder | 3.53 | 2.53 | 4.92 | 1.1E-13 | 1.3E-13 |
| Personality disorder | 8.65 | 6.69 | 11.20 | 1.9E-60 | 7.6E-60 |

#### **Table S5** Sex differences in the association between adolescent anxiety, risk stratifiers and adult-onset psychiatric disorders.

#### Sex stratified results are presented, and interaction results refer to the interaction between sex and stratifier.

| **Risk stratifier** | **Females** | | | | | **Males** | | | | | | | | **Interaction** | | | | | | |
| --- | --- | --- | --- | --- | --- | --- | --- | --- | --- | --- | --- | --- | --- | --- | --- | --- | --- | --- | --- | --- |
|  | **Hazard ratio** | **95% CI** | | **P-value** | | **Hazard ratio** | | **95% CI** | | | | **P-value** | | **Hazard ratio** | | **95% CI** | | | | **P-value** |
|  |  | **Lower** | **Upper** |  |  |  |  | **Lower** | | **Upper** | |  |  |  |  | **Lower** | | **Upper** | |  |
| Adolescent anxiety | 3.26 | 2.87 | 3.72 | 1.3E-71 | | 3.48 | | 2.64 | | 4.58 | | 7.3E-19 | | 0.97 | | 0.70 | | 1.35 | | 0.87 |
| ***Polygenic scores*** | | | | |  | |  | |  | |  | |  | |  | |  | |  | |
| ADHD PGS | 1.12 | 0.97 | 1.30 | 0.11 | | 1.54 | | 1.10 | | 2.14 | | 0.011 | | 0.81 | | 0.55 | | 1.20 | | 0.29 |
| Anxiety PGS | 1.10 | 0.95 | 1.28 | 0.19 | | 1.29 | | 0.95 | | 1.76 | | 0.10 | | 0.86 | | 0.59 | | 1.25 | | 0.43 |
| Bipolar disorder PGS | 1.10 | 0.95 | 1.27 | 0.19 | | 1.16 | | 0.84 | | 1.61 | | 0.36 | | 0.99 | | 0.67 | | 1.46 | | 0.96 |
| MDD PGS | 1.08 | 0.93 | 1.25 | 0.31 | | 1.46 | | 1.08 | | 1.98 | | 0.01 | | 0.75 | | 0.52 | | 1.08 | | 0.12 |
| OCD PGS | 1.00 | 0.87 | 1.16 | 0.95 | | 1.48 | | 1.07 | | 2.05 | | 0.02 | | 0.61 | | 0.41 | | 0.90 | | 0.013 |
| PTSD PGS | 1.19 | 1.02 | 1.38 | 0.023 | | 1.42 | | 1.03 | | 1.96 | | 0.03 | | 0.82 | | 0.56 | | 1.20 | | 0.30 |
| Schizophrenia PGS | 1.00 | 0.86 | 1.15 | 0.97 | | 1.21 | | 0.90 | | 1.61 | | 0.21 | | 0.76 | | 0.53 | | 1.08 | | 0.13 |
| ***Parental history*** | | | | |  | |  | |  | |  | |  | |  | |  | |  | |
| Parental history of any psychiatric disorder | 1.12 | 0.87 | 1.43 | 0.38 | | 0.90 | | 0.52 | | 1.56 | | 0.71 | | 0.94 | | 0.49 | | 1.78 | | 0.84 |
| Parental history of severe mental illness | 2.20 | 1.43 | 3.37 | 3.0E-04 | | NA – insufficient cell count | | | | | | | | NA | | | | | | |
| Parental history of anxiety or depression | 1.13 | 0.87 | 1.46 | 0.35 | | 0.60 | | 0.32 | | 1.11 | | 0.10 | | 1.45 | | 0.72 | | 2.92 | | 0.30 |
| ***Demographic and clinical factors*** | | | | |  | |  | |  | |  | |  | |  | |  | |  | |
| Psychiatric co-morbidity | 1.07 | 0.84 | 1.36 | 0.57 | | 1.77 | | 1.03 | | 3.06 | | 0.039 | | 0.49 | | 0.26 | | 0.93 | | 0.028 |
| Neurodevelopmental comorbidity | 0.90 | 0.58 | 1.37 | 0.61 | | NA – insufficient cell count | | | | | | | | NA | | | | | | |

#### **Table S6**. Hazard ratios of the likelihood of experiencing an adult-onset psychiatric disorder (≥18 years) by each predictor in the full MoBa sample.

Results of the interaction between adolescent anxiety diagnosis and stratifier are also presented.

| **Predictor** | **Whole sample** | | | | **Interaction with anxiety diagnosis** | | | |
| --- | --- | --- | --- | --- | --- | --- | --- | --- |
|  | **Hazard ratio** | **95% CI** | | **P-value** | **Hazard ratio** | **95% CI** | | **P-value** |
|  |  | **Lower** | **Upper** |  |  | **Lower** | **Upper** |  |
| ***Polygenic scores*** | | | | |  |  |  |  |
| ADHD PGS | 1.24 | 1.17 | 1.30 | 1.0E-15 | 0.91 | 0.61 | 1.36 | 0.64 |
| Anxiety PGS | 1.27 | 1.20 | 1.34 | 2.4E-19 | 1.02 | 0.68 | 1.54 | 0.91 |
| Bipolar disorder PGS | 1.20 | 1.14 | 1.27 | 3.0E-12 | 0.90 | 0.63 | 1.29 | 0.57 |
| MDD PGS | 1.42 | 1.35 | 1.49 | 2.1E-39 | 1.04 | 0.70 | 1.55 | 0.85 |
| OCD PGS | 1.03 | 0.98 | 1.08 | 0.27 | 1.12 | 0.75 | 1.67 | 0.58 |
| PTSD PGS | 1.32 | 1.25 | 1.39 | 6.1E-26 | 0.96 | 0.64 | 1.44 | 0.83 |
| Schizophrenia PGS | 1.12 | 1.06 | 1.18 | 1.6E-05 | 0.84 | 0.57 | 1.24 | 0.38 |
| ***Parental history*** | | | | |  |  |  |  |
| Parental history of any psychiatric disorder | 1.72 | 1.57 | 1.89 | 4.2E-31 | 0.51 | 0.24 | 1.06 | 0.072 |
| Parental history of severe mental illness | 2.50 | 2.07 | 3.01 | 6.0E-22 | 1.71 | 0.63 | 4.63 | 0.29 |
| Parental history of anxiety or depression | 1.61 | 1.46 | 1.77 | 2.5E-22 | 0.47 | 0.21 | 1.03 | 0.058 |
| ***Demographic and clinical factors*** | | | | |  |  |  |  |
| Psychiatric co-morbidity | 3.45 | 3.07 | 3.86 | 9.4E-100 | 0.52 | 0.24 | 1.15 | 0.11 |
| Neurodevelopmental comorbidity | 1.69 | 1.41 | 2.03 | 1.8E-08 | 0.22 | 0.05 | 1.03 | 0.054 |
| Female sex | 2.62 | 2.38 | 2.88 | 4.1E-89 | 0.80 | 0.37 | 1.73 | 0.57 |

#### **Table S7** Hazard ratios of the association between polygenic scores (PGS) and specific categories of psychiatric outcomes in individuals with adolescent anxiety.

CI= 95% confidence interval. MDD=major depressive disorder. OCD=obsessive compulsive disorder. PTSD=post-traumatic stress disorder.

| **Outcome** | **PGS** | **Hazard Ratio** | **Lower CI** | **Upper CI** | **P-value** |
| --- | --- | --- | --- | --- | --- |
| Psychosis | ADHD | 1.07 | 0.55 | 2.07 | 0.84 |
|  | Anxiety | 2.53 | 1.34 | 4.76 | 4.0E-03 |
|  | Bipolar disorder | 2.60 | 1.27 | 5.32 | 0.01 |
|  | MDD | 2.60 | 1.35 | 5.03 | 4.5E-03 |
|  | OCD | 1.14 | 0.60 | 2.19 | 0.69 |
|  | PTSD | 1.31 | 0.67 | 2.56 | 0.44 |
|  | Schizophrenia | 1.14 | 0.60 | 2.15 | 0.69 |
| Bipolar disorder | ADHD | 1.58 | 1.03 | 2.42 | 0.03 |
|  | Anxiety | 0.96 | 0.63 | 1.46 | 0.85 |
|  | Bipolar disorder | 1.32 | 0.86 | 2.02 | 0.20 |
|  | MDD | 1.24 | 0.82 | 1.87 | 0.31 |
|  | OCD | 1.07 | 0.70 | 1.62 | 0.77 |
|  | PTSD | 1.55 | 1.01 | 2.38 | 0.05 |
|  | Schizophrenia | 1.11 | 0.74 | 1.67 | 0.62 |
| MDD | ADHD | 1.07 | 0.85 | 1.34 | 0.57 |
|  | Anxiety | 1.12 | 0.89 | 1.40 | 0.34 |
|  | Bipolar disorder | 1.05 | 0.84 | 1.32 | 0.67 |
|  | MDD | 0.98 | 0.78 | 1.23 | 0.86 |
|  | OCD | 1.27 | 1.01 | 1.59 | 0.04 |
|  | PTSD | 1.16 | 0.92 | 1.46 | 0.20 |
|  | Schizophrenia | 1.06 | 0.85 | 1.32 | 0.60 |
| OCD | ADHD | 1.38 | 0.86 | 2.20 | 0.18 |
|  | Anxiety | 1.19 | 0.75 | 1.90 | 0.46 |
|  | Bipolar disorder | 0.89 | 0.56 | 1.40 | 0.60 |
|  | MDD | 0.87 | 0.55 | 1.37 | 0.55 |
|  | OCD | 1.32 | 0.83 | 2.11 | 0.23 |
|  | PTSD | 1.02 | 0.64 | 1.63 | 0.94 |
|  | Schizophrenia | 1.14 | 0.73 | 1.79 | 0.56 |
| PTSD | ADHD | 1.24 | 0.89 | 1.74 | 0.20 |
|  | Anxiety | 1.20 | 0.86 | 1.68 | 0.27 |
|  | Bipolar disorder | 1.15 | 0.82 | 1.60 | 0.42 |
|  | MDD | 1.27 | 0.92 | 1.77 | 0.15 |
|  | OCD | 1.08 | 0.77 | 1.50 | 0.67 |
|  | PTSD | 1.24 | 0.89 | 1.75 | 0.21 |
|  | Schizophrenia | 1.29 | 0.93 | 1.78 | 0.13 |
| Eating disorder | ADHD | 1.12 | 0.78 | 1.62 | 0.53 |
|  | Anxiety | 0.84 | 0.59 | 1.21 | 0.35 |
|  | Bipolar disorder | 1.30 | 0.90 | 1.87 | 0.16 |
|  | MDD | 1.09 | 0.76 | 1.55 | 0.64 |
|  | OCD | 1.38 | 0.96 | 1.97 | 0.08 |
|  | PTSD | 1.05 | 0.73 | 1.52 | 0.78 |
|  | Schizophrenia | 1.10 | 0.77 | 1.55 | 0.61 |
| Personality disorder | ADHD | 1.34 | 1.01 | 1.77 | 0.04 |
|  | Anxiety | 1.17 | 0.89 | 1.55 | 0.27 |
|  | Bipolar disorder | 1.13 | 0.85 | 1.49 | 0.40 |
|  | MDD | 1.12 | 0.85 | 1.47 | 0.42 |
|  | OCD | 1.05 | 0.79 | 1.38 | 0.75 |
|  | PTSD | 1.23 | 0.93 | 1.63 | 0.15 |
|  | Schizophrenia | 0.90 | 0.69 | 1.18 | 0.44 |

### Supplementary Figures

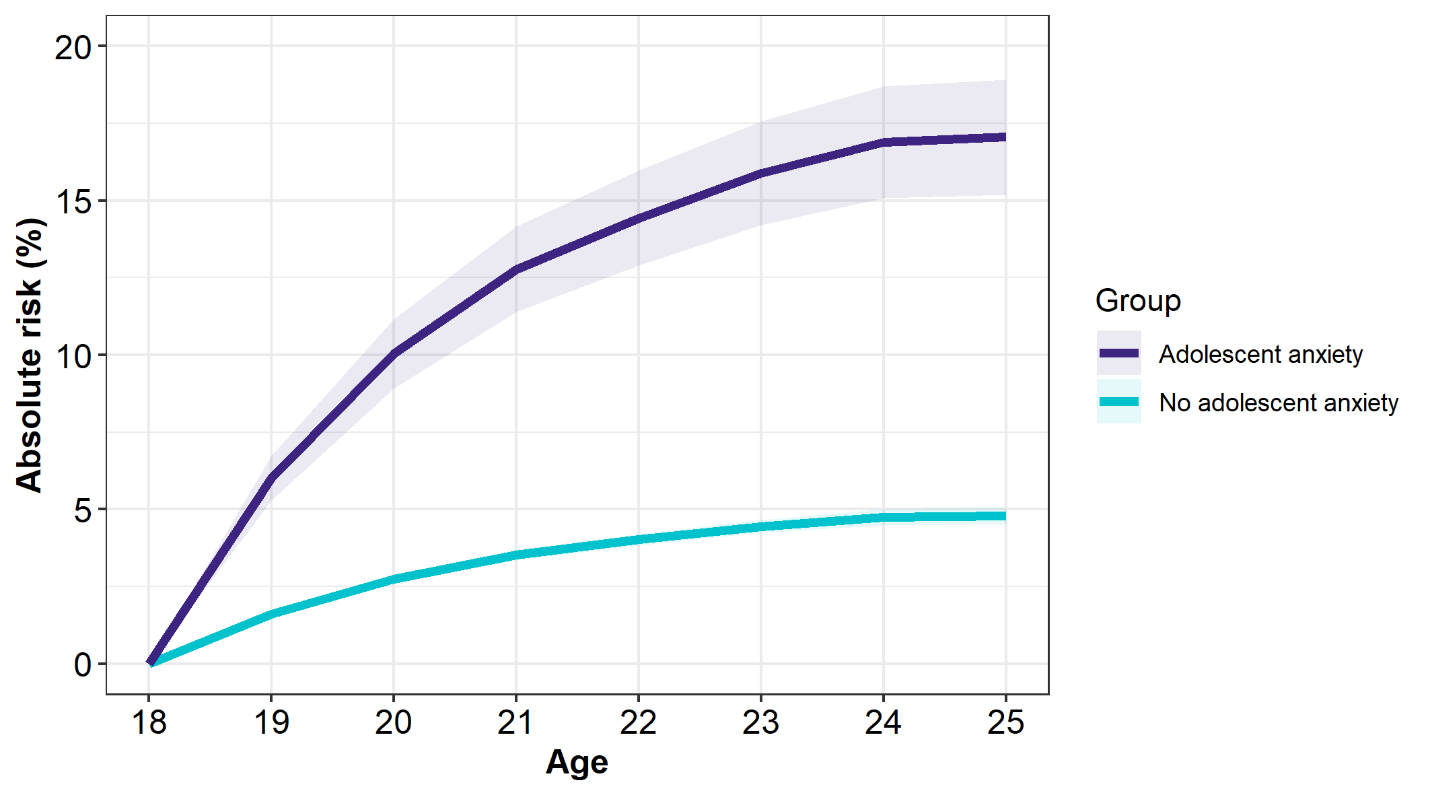

**Figure S1.** Absolute risk of developing a young adult-onset psychiatric disorder, stratified by adolescent anxiety status. Purple refers to individuals with adolescent anxiety, blue refers to individuals without adolescent anxiety.

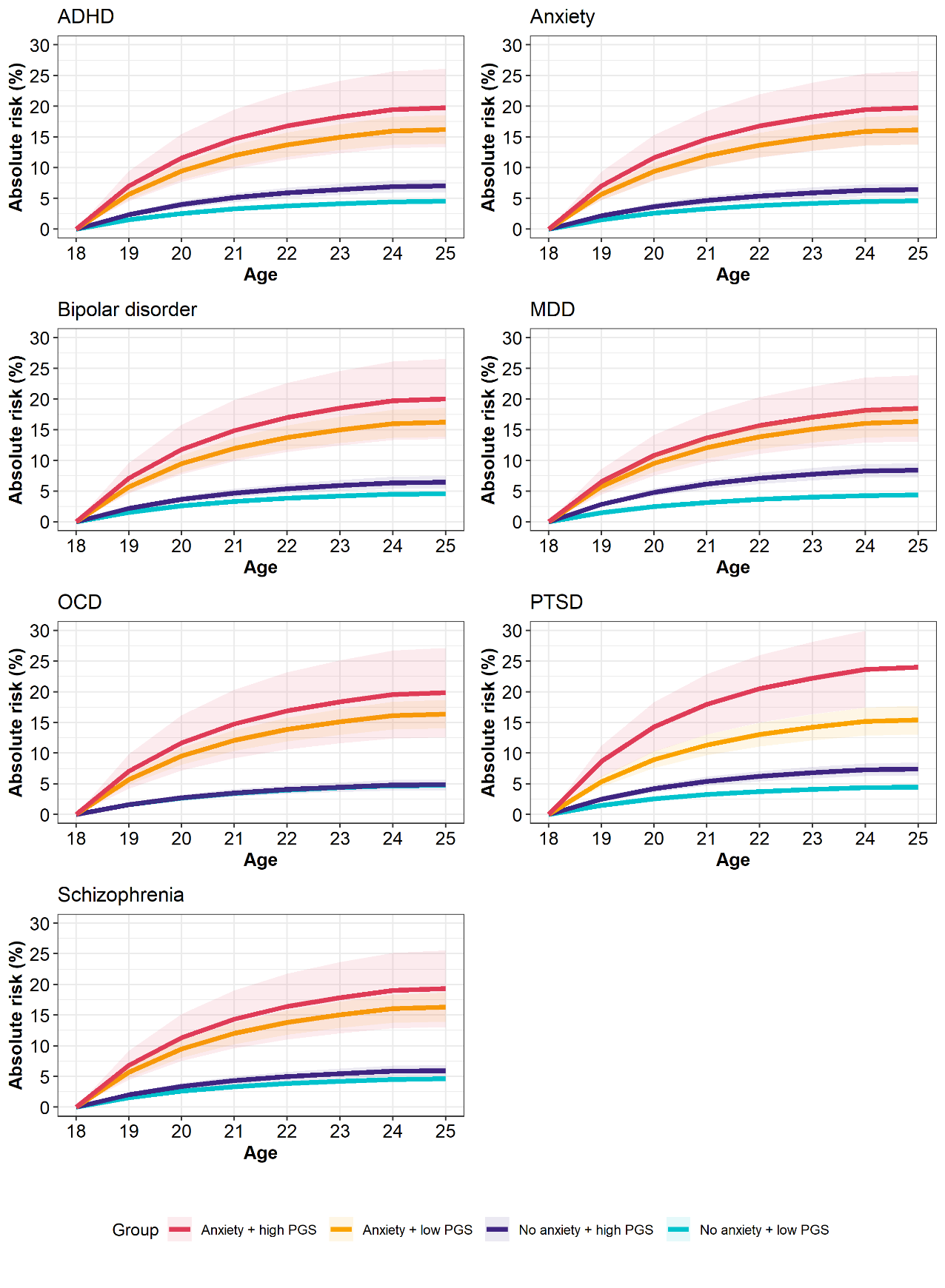

**Figure S3.** Absolute risk of an adult-onset psychiatric disorder, stratified by polygenic score (PGS) and adolescent anxiety status. PGS is indicated by the title of each individual plot. ADHD – Attention deficit hyperactivity disorder; MDD – major depressive disorder; OCD – obsessive compulsive disorder; PTSD – post-traumatic stress disorder.
